# Burden of intraventricular haemorrhage among preterm neonates in Tanzania: A prospective observational study

**DOI:** 10.64898/2025.12.06.25339943

**Authors:** Immaculate B. Ndawi, Neema Passian Chami, Patrick S. Ngoya

## Abstract

**Background:** Intraventricular haemorrhage (IVH) is a common intracranial complication of preterm neonates, especially those born before 32 weeks of gestation, and poses a high risk for morbidity with lifelong neurological disability. IVH remains a dilemma in low-resource settings, where access to high-quality prenatal and neonatal care is very limited. Cranial ultrasound (USS) remains an accessible imaging tool for detection of IVH. This study aimed to determine the burden and factors associated with IVH among preterm neonates admitted at a tertiary hospital in Tanzania.

**Methodology:** This was a prospective observational study on admitted preterm neonates born at gestational age (GA) of less than 37 weeks from March 2024 to February 2025. Cranial USS was performed to detect and grade IVH using the Papile Classification. Univariate and multivariate logistic regression was used to determine factors associated with IVH.

**Results:** IVH was prevalent in 96 of the 410 preterm neonates (23.4%). Majority (69.8%) presented with IVH grade I. Level of agreement for IVH grading was near perfect between the two reviewers. Independent risk factors for IVH on multivariate analysis were very low birth weight (VLBW) (aOR=7.2, 95%CI=2.1-24.8), extremely low birth weight (ELBW) (aOR= 6.3, 95%CI=1.7-23.1), respiratory distress syndrome (RDS) (aOR=2.3, 95%CI= 1.2-4.6), preterm premature rupture of membranes (PPROM) (aOR=2.3, 95%CI=1.2-4.4).

**Conclusions:** There is still a significant IVH burden with low grade IVH predominating in Tanzania. Modifiable risk factors associated with IVH included RDS and PPROM which can be potentially be targeted with interventions to reduce IVH.

## INTRODUCTION

Intraventricular haemorrhage (IVH) is a common intracranial complication of preterm neonates, especially those born before 32 weeks of gestation, and poses a high risk for morbidity with lifelong neurological disability [1,2] The etiology of IVH is multifactorial and is primarily attributed to the intrinsic fragility of the richly vascularized germinal matrix and the disturbance in the cerebral blood flow [3]. It is well established that in the transition from fetal to neonatal circulation, cerebral autoregulation is severely impaired in preterm neonates which increases the risk of IVH development especially within 72 hours of life of a preterm neonate [4].

The pooled prevalence of IVH among preterm neonates in Africa and especially in sub-saharan Africa has been reported to be around 29% [5,6]. There is paucity of data on the prevalence of IVH among preterm neonates in Tanzania, with a previous study demonstrating a 62% IVH rate [7] With advancement in medical technology and treatment, there has been a decline in incidence of IVH in developed countries [8]. Despite such improvements, IVH remains a major challenge in low resource settings, where access to high-quality prenatal and neonatal care is very limited. Cranial USS remains a portable non-invasive, dynamic imaging tool for detection of IVH. Therefore, this study aimed to determine the burden and factors associated with IVH among preterm neonates admitted at a tertiary hospital in Tanzania.

## MATERIALS AND METHODS

### Study design, duration and setting

This was a hospital-based, prospective observational study that was carried out from March 2024 to February 2025 at Bugando Medical Centre (BMC), a tertiary hospital in Mwanza, Tanzania.

### Study population

All preterm neonates admitted between 1^st^ March 2024 to 31^st^ July 2024 at the neonatal unit were recruited.

### Eligibility criteria

All preterm neonates admitted at the neonatal care unit of BMC, born at gestational age (GA) of less than 37 weeks were included. Preterm neonates with scalp, skull or brain malformations that obscured detection of IVH were excluded.

### Study variables

Exposure variables included maternal age, parity, antenatal clinic visits, antenatal steroids use, hypertension, diabetes mellitus, infection, HIV status, preterm premature rupture of membranes (PPROM) as well as neonatal gestational age at birth, birth weight, gender, mode of delivery, resuscitation, convulsions, presence of pallor, cyanosis, jaundice or hypothermia, haemoglobin (Hb) and platelet level, ventilation required, respiratory distress syndrome (RDS) and surfactant therapy use. Outcome variable was presence of IVH.

### Sample size

A minimum sample size of 363 participants, with an additional 10% to account for participant loss, was calculated using the Kish formula with a confidence level of 95%, a precision level of 5% and assuming an IVH prevalence of 62% in Tanzania [7,9]. Consecutive sampling technique was employed.

### Data collection and procedures

A study participants’ checklist was used to collect data. Gestation age at birth was recorded and later categorized into late preterm (32 to less than 37 weeks), very preterm (28 to less than 32 weeks), and extremely preterm (less than 28 weeks). Birth weight in grams (g) was categorized as low birth weight (LBW) (1500 to less than 2500g), very LBW (VLBW) (1000 to less than 1500g), extremely LBW (ELBW) (less than 1000g). Presence of hypothermia if temperature less than 36.5 degrees Celsius. Haemoglobin and platelets level was measured in grams per deciliter (g/dl) and x10^9^/Litre (L) respectively. Neonates were followed up over a period of 7 days to detect IVH by cranial USS using a portable ultrasound Sonosite M-Turbo (Fujifilm, Japan). IVH was graded into grade I-IV by two reviewers using the Papile classification [10].

### Data management and analysis

Data was collected from the participants’ checklists was entered into Microsoft Excel (Microsoft Corporation, USA) and then exported to Stata version 17 (StataCorp LLC, USA) for cleaning and analysis. Data was presented as frequencies or proportions for categorical variables. Cohen’s kappa statistics were used to assess the level of agreement between the two separate reviewers, one radiologist and one neonatologist, blinded to baseline maternal and neonatal characteristics. Univariate and multivariate logistic regression was applied to determine risk factors associated with IVH. A p-value of <0.05 was considered statistically significant.

## RESULTS

### Baseline maternal and neonatal characteristics

Upto 410 out of 464 admitted preterm neonates were enrolled into the study. 54 of the admitted preterm neonates were excluded due to various reasons as shown on **Figure 1**.

**Fig. 1.**
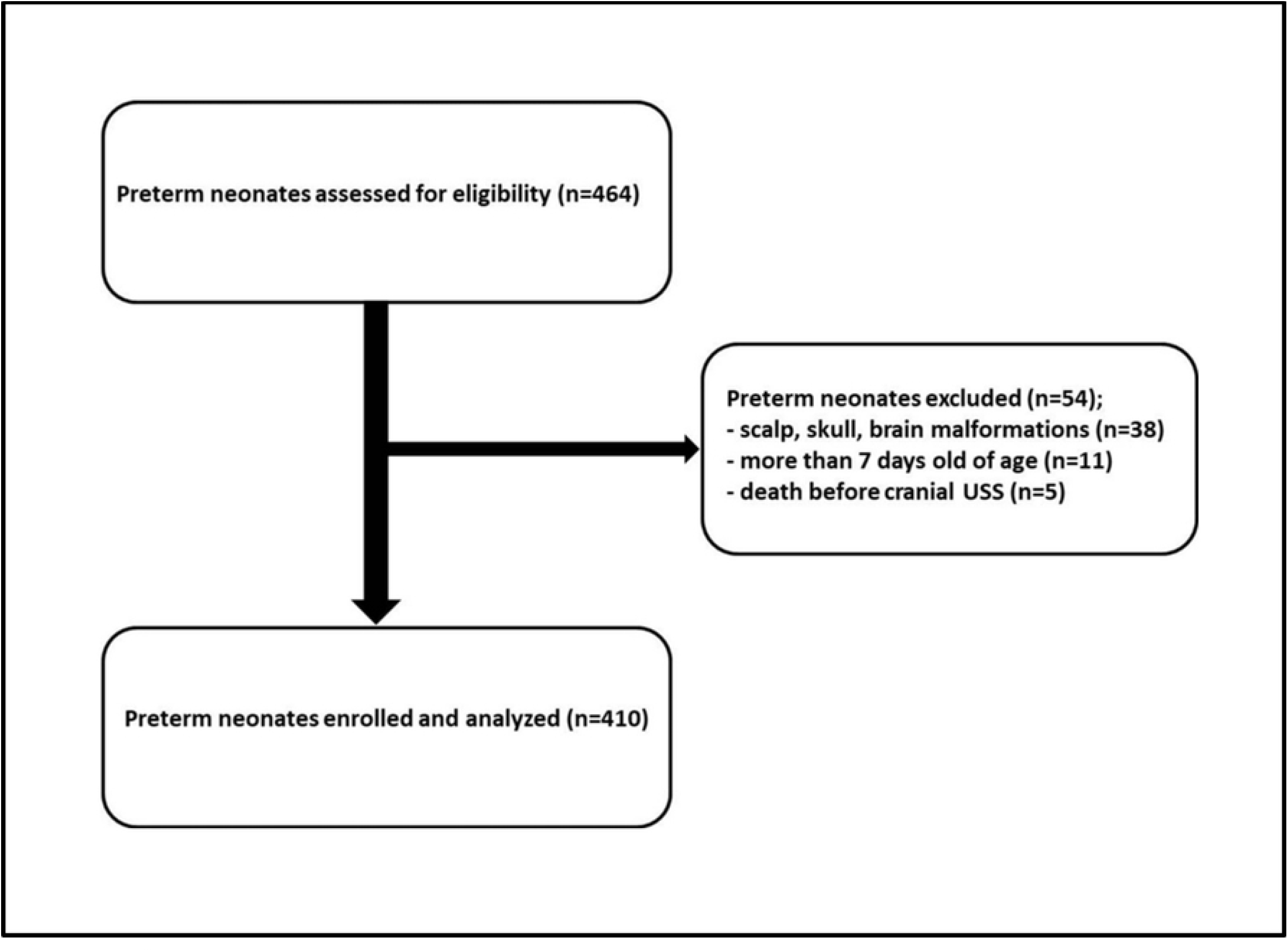

Majority of the mothers of the preterm neonates were aged between 19 and 34 years (72%), having used antenatal steroids (79%), and had experienced PPROM (73%). About 4 - 22% mothers had underlying comorbidities as shown in **Table 1**.

**Table 1.** Baseline maternal characteristics (n=410)

| Variable | Category | n (%) |
| --- | --- | --- |
| Age, years | <18 | 34 (8.3) |
|  | 19-34 | 295 (72.0) |
|  | >35 | 81 (19.7) |
| Parity | 1 | 303 (74.0) |
|  | 2 or more | 107 (26.0) |
| ANC visits | No | 63 (15.0) |
|  | Yes | 347 (85.0) |
| Antenatal steroids | No | 87 (21.0) |
|  | Yes | 323 (79.0) |
| Hypertension | No | 319 (78.0) |
|  | Yes | 91 (22.0) |
| Diabetes mellitus | No | 394 (96.1) |
|  | Yes | 16 (3.9) |
| Infection | No | 347 (85.0) |
|  | Yes | 63 (15.0) |
| HIV positive | No | 395 (96.3) |
|  | Yes | 15 (3.7) |
| PPROM | No | 111 (27.0) |
|  | Yes | 299 (73.0) |

ANC, antenatal clinic; HIV, human immune-deficiency virus; PPRM, preterm premature rupture of membranes.

Most of the preterm neonates recruited were very preterm (28 - <32 weeks) and male: female ratio of 1:1.3. Many of the preterm neonates presented with hypothermia (57.3%), required ventilation (94.9%), and were affected with RDS (76.3%) (**Table 2**).

**Table 2.**
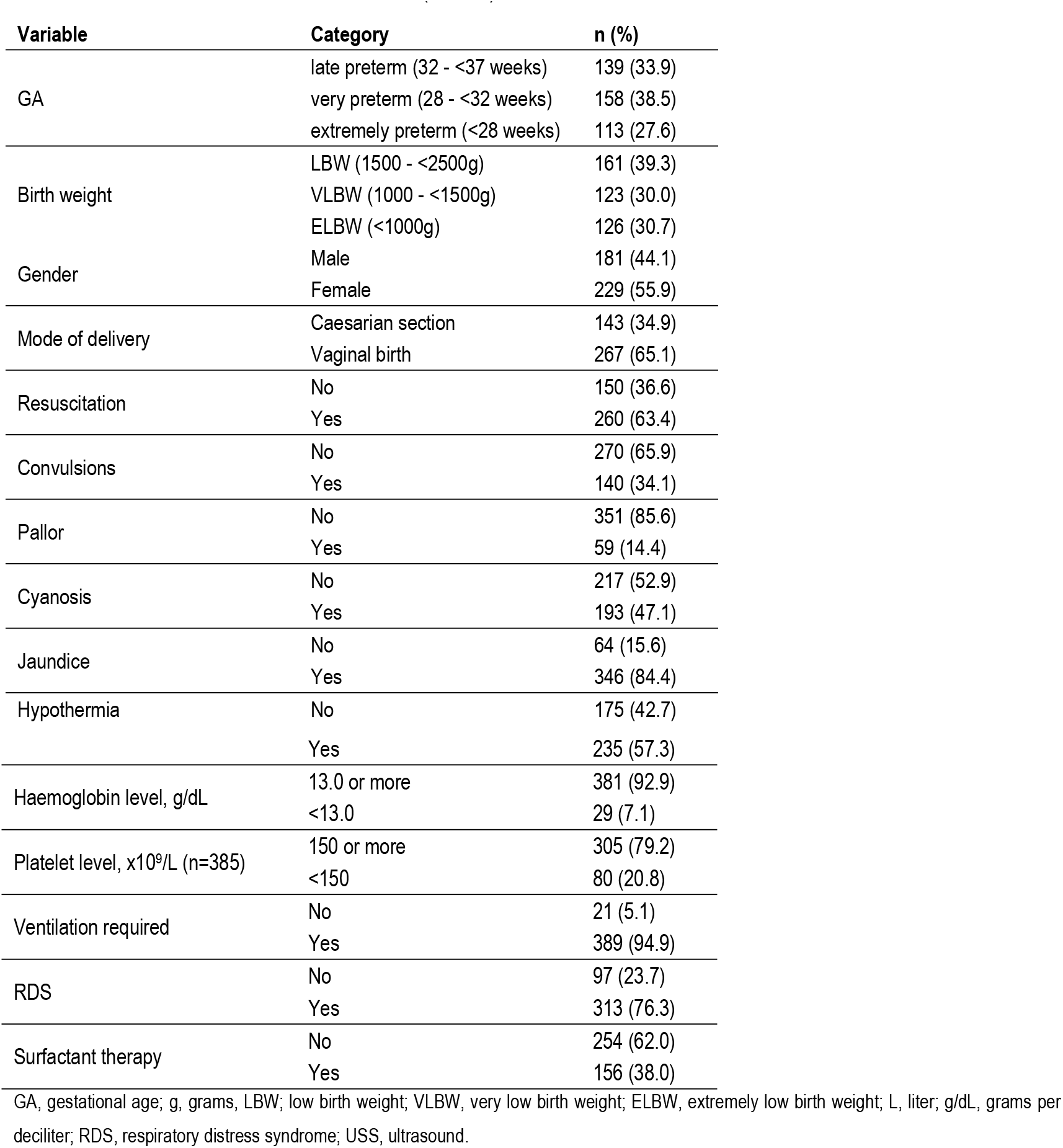
Baseline neonatal characteristics (n=410)

### Prevalence of IVH

IVH was detected in 96 out of the 410 (23.4%) preterm neonates. IVH was graded into 4 grades by cranial USS (**Figure 2**). Of those who had IVH, majority (69.8%) had IVH grade I (**Figure 3**). Low grade IVH (grade I and II combined) constituted 81% while 19% had high grade IVH (grade III and IV combined).

**Fig. 2.**
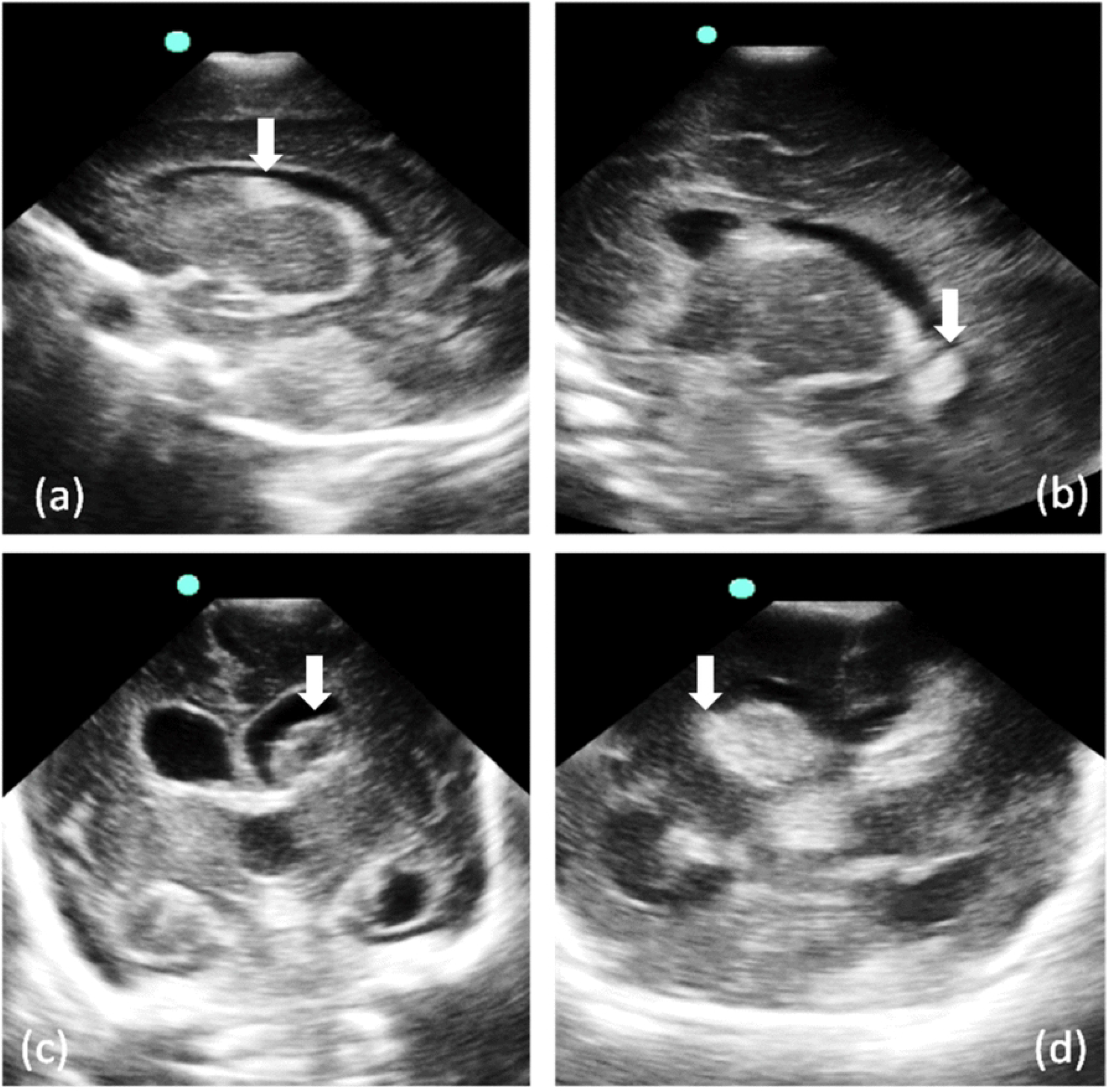

**Fig. 3.**
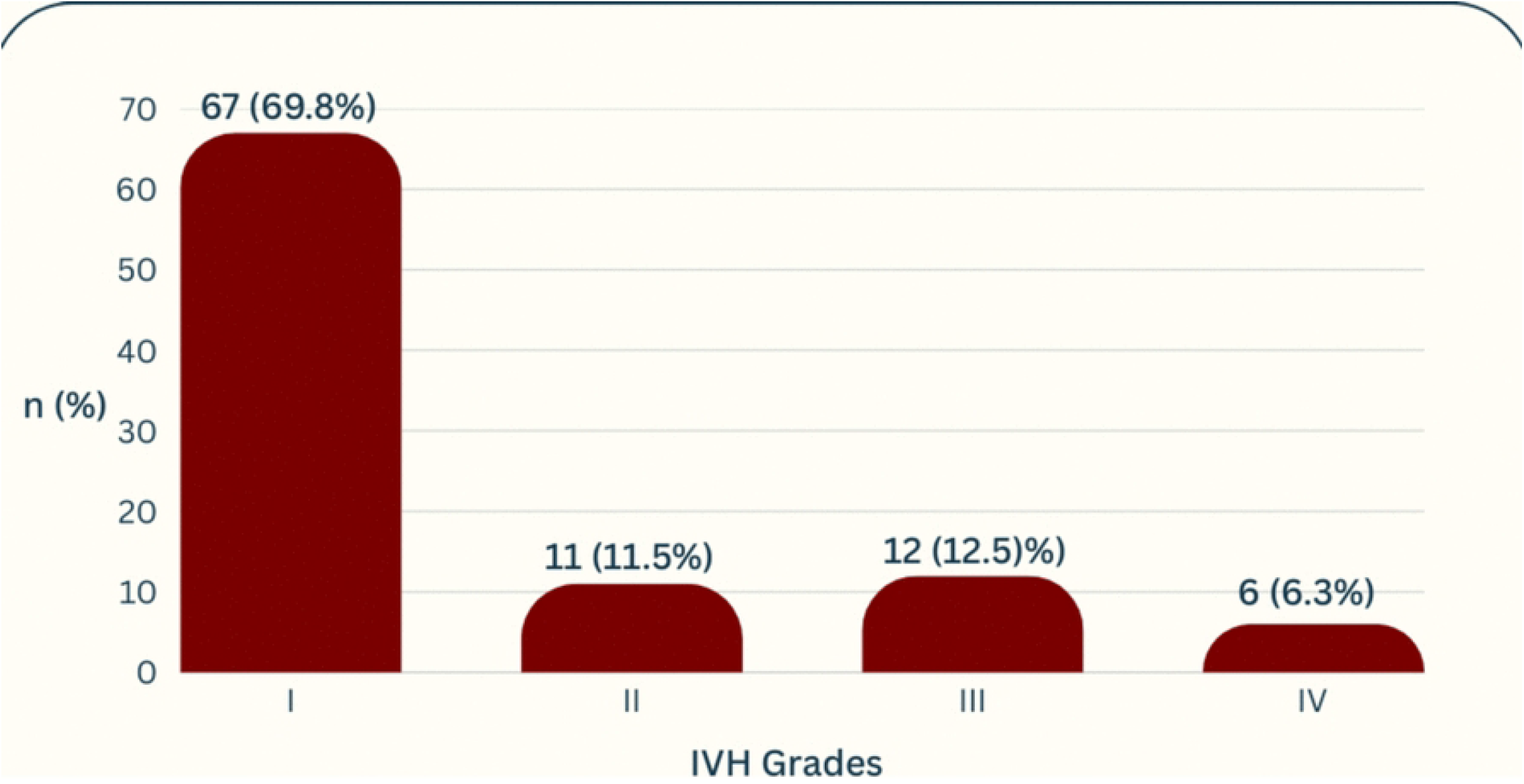

### Level of agreement in the detection of IVH

There was a 98.3% near perfect agreement in the detection of IVH grades between the two reviewers by Cohen’s kappa statistics (expected agreement=64.0%, kappa=0.95, z-score=19.3, p<0.001).

### Factors associated with IVH

Independent significant risk factors for IVH on multivariate analysis were VLBW (aOR=7.2, 95%CI=2.1-24.8), ELBW (aOR=6.3, 95%CI=1.7-23.1), RDS (aOR=2.3, 95%CI= 1.2-4.6), PPROM (aOR=2.3, 95%CI= 1.2-4.4), hypothermia (aOR=0.4, 95%CI=0.2-0.6) and cyanosis (aOR=0.4, 95%CI= 0.2-1.0), as shown in **Table 3 and Table 4** and as illustrated in **Figure 4**.

**Fig. 4.**
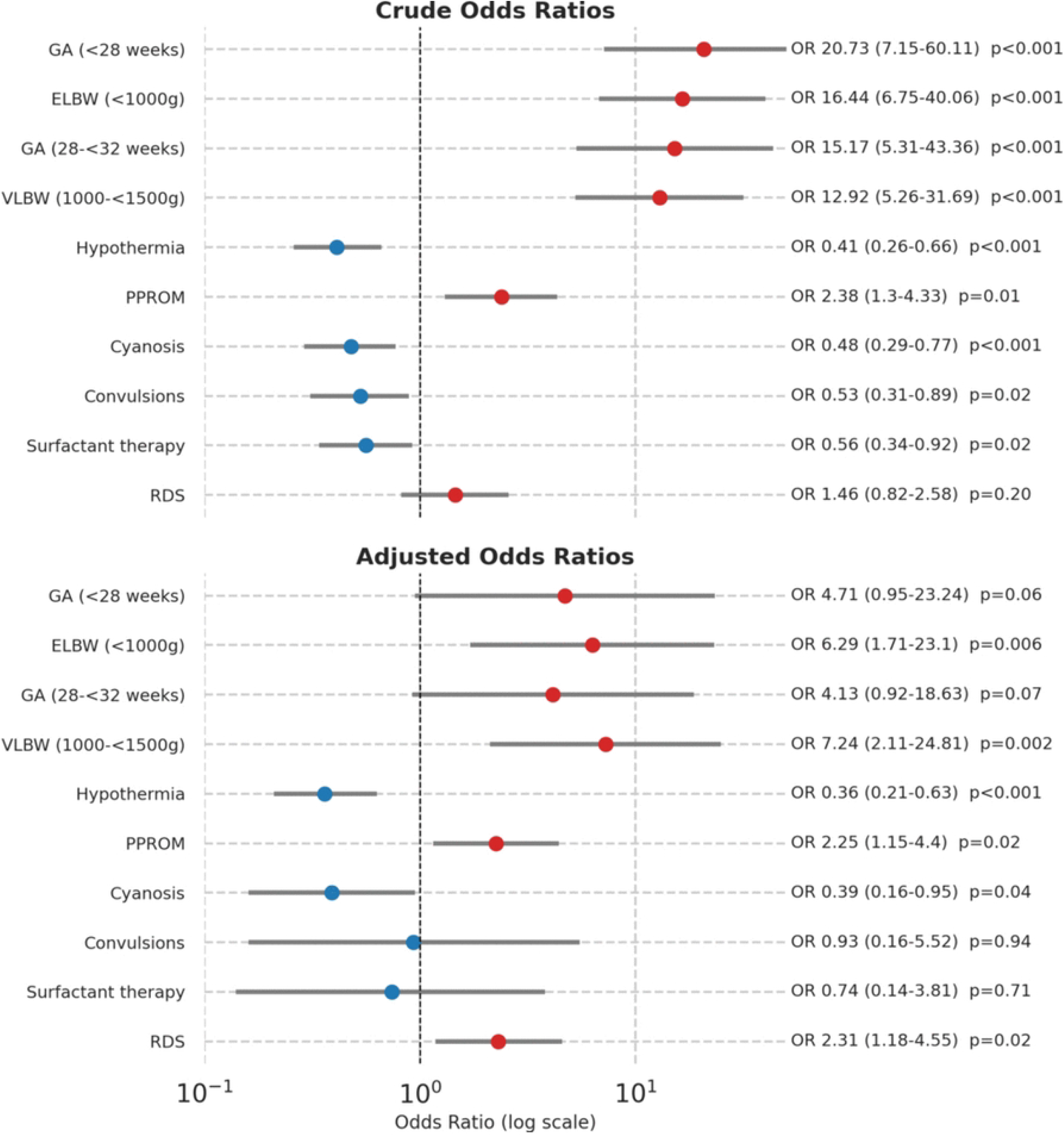

**Table 3.**
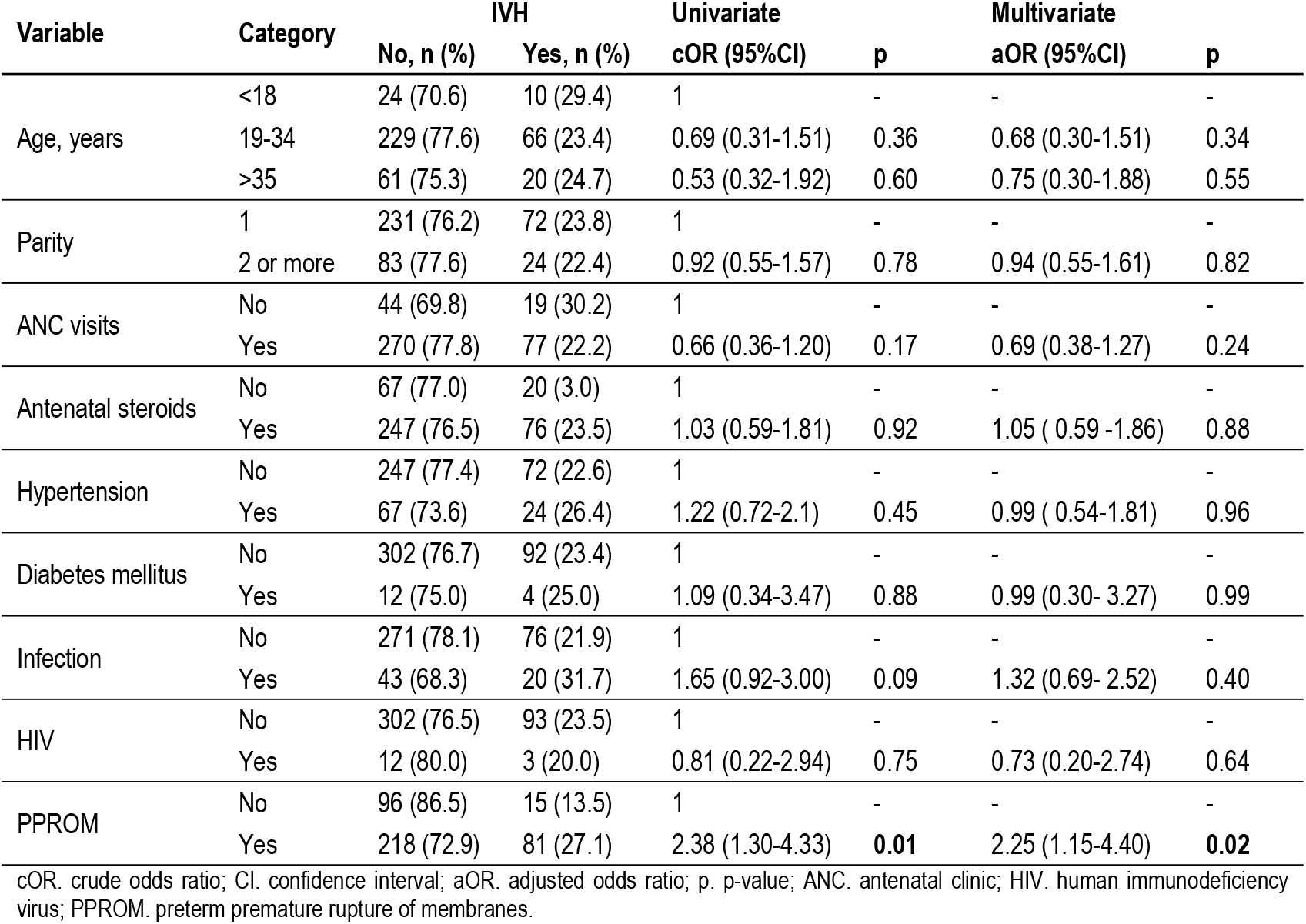
Maternal factors associated with IVH.

| Variable | Category | IVH |  | Univariate |  | Multivariate |  |
| --- | --- | --- | --- | --- | --- | --- | --- |
|  |  | No, n (%) | Yes, n (%) | cOR (95%CI) | p | aOR (95%CI) | p |
| Age, years | <18 | 24 (70.6) | 10 (29.4) | 1 | - | - | - |
|  | 19-34 | 229 (77.6) | 66 (23.4) | 0.69 (0.31-1.51) | 0.36 | 0.68 (0.30-1.51) | 0.34 |
|  | >35 | 61 (75.3) | 20 (24.7) | 0.53 (0.32-1.92) | 0.60 | 0.75 (0.30-1.88) | 0.55 |
| Parity | 1 | 231 (76.2) | 72 (23.8) | 1 | - | - | - |
|  | 2 or more | 83 (77.6) | 24 (22.4) | 0.92 (0.55-1.57) | 0.78 | 0.94 (0.55-1.61) | 0.82 |
| ANC visits | No | 44 (69.8) | 19 (30.2) | 1 | - | - | - |
|  | Yes | 270 (77.8) | 77 (22.2) | 0.66 (0.36-1.20) | 0.17 | 0.69 (0.38-1.27) | 0.24 |
| Antenatal steroids | No | 67 (77.0) | 20 (3.0) | 1 | - | - | - |
|  | Yes | 247 (76.5) | 76 (23.5) | 1.03 (0.59-1.81) | 0.92 | 1.05 ( 0.59 -1.86) | 0.88 |
| Hypertension | No | 247 (77.4) | 72 (22.6) | 1 | - | - | - |
|  | Yes | 67 (73.6) | 24 (26.4) | 1.22 (0.72-2.1) | 0.45 | 0.99 ( 0.54-1.81) | 0.96 |
| Diabetes mellitus | No | 302 (76.7) | 92 (23.4) | 1 | - | - | - |
|  | Yes | 12 (75.0) | 4 (25.0) | 1.09 (0.34-3.47) | 0.88 | 0.99 (0.30- 3.27) | 0.99 |
| Infection | No | 271 (78.1) | 76 (21.9) | 1 | - | - | - |
|  | Yes | 43 (68.3) | 20 (31.7) | 1.65 (0.92-3.00) | 0.09 | 1.32 (0.69- 2.52) | 0.40 |
| HIV | No | 302 (76.5) | 93 (23.5) | 1 | - | - | - |
|  | Yes | 12 (80.0) | 3 (20.0) | 0.81 (0.22-2.94) | 0.75 | 0.73 (0.20-2.74) | 0.64 |
| PPROM | No | 96 (86.5) | 15 (13.5) | 1 | - | - | - |
|  | Yes | 218 (72.9) | 81 (27.1) | 2.38 (1.30-4.33) | **0.01** | 2.25 (1.15-4.40) | **0.02** |

cOR. crude odds ratio; CI. confidence interval; aOR. adjusted odds ratio; p. p-value; ANC. antenatal clinic; HIV. human immunodeficiency virus; PPRM. preterm premature rupture of membranes.

**Table 4.**
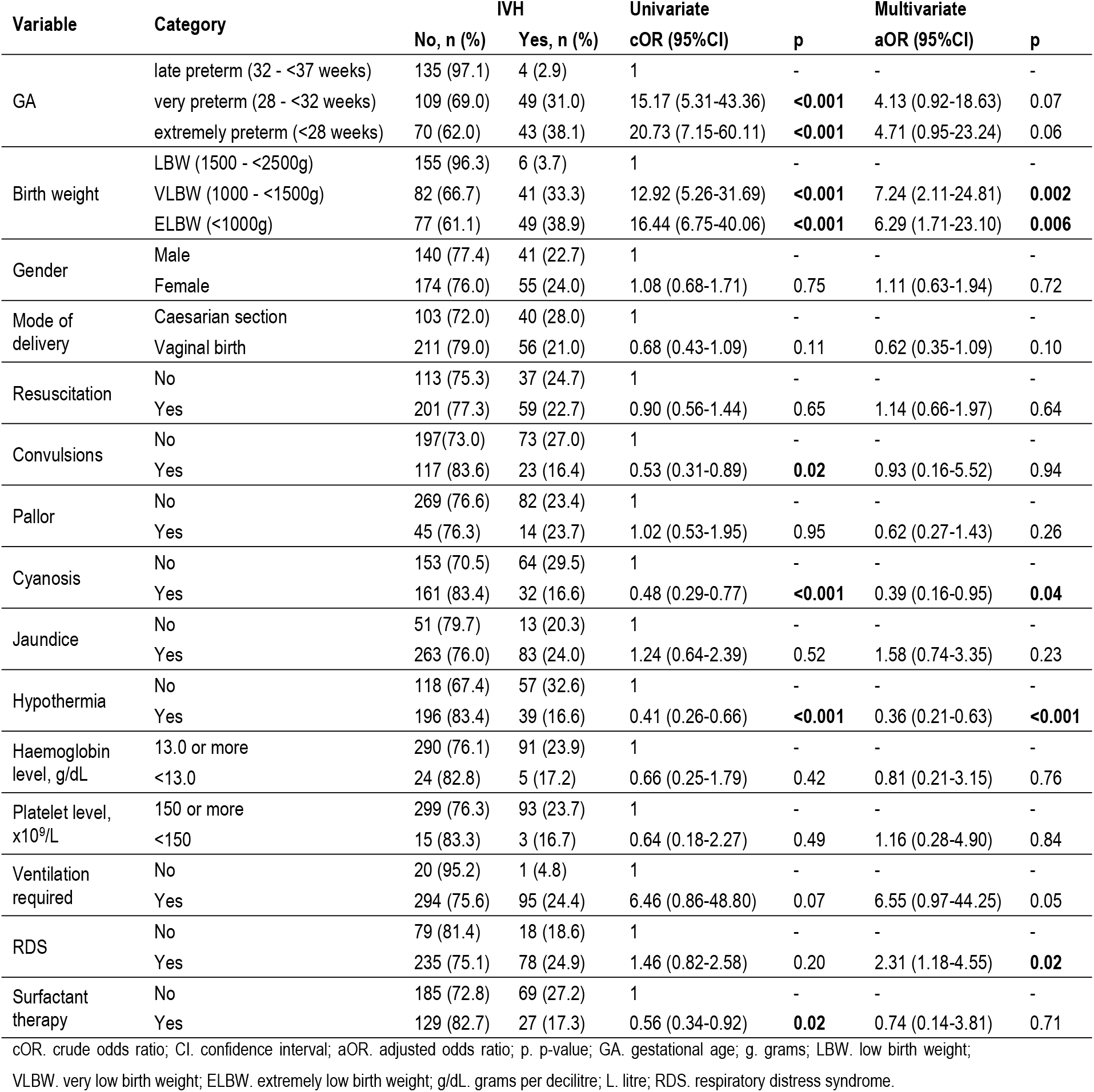
Neonatal factors associated with IVH.

## DISCUSSION

The IVH prevalence observed in this study (23.4%) markedly contrasts with a prior study conducted in a comparable Tanzanian hospital setting, which reported a higher IVH rate of 62% [7] Our finding is almost identical to a South Africa study which showed an IVH prevalence of 22.6% [11]. Additionally, the pooled African IVH prevalence estimated at 29% is slightly higher [5,6] These differences may be attributed to heterogeneities in the study design, methodology, or population characteristics.

Our findings, contribute to a growing body of evidence suggesting that advancements in medical technology and neonatal care have contributed to a reduction in IVH prevalence (from 62% to 23%) in a low- and middle-income country such as Tanzania. This is apparently dissimilar to a recent whole-population cohort study that reported an increase in IVH rates despite advances in neonatal care in developed countries, an observation for which the underlying causes remain unclear [12].

Low grade IVH (grade I or II) constituted 81% in our study. Previously, low grade IVH was not considered to have clinically significant neurodevelopmental impairment compared to high grade IVH [13] A recent systematic review and meta-analysis whose primary outcome was to determine neurodevelopmental impairment after 5 years of age, has shown that low-grade IVH is associated with a lower intelligence quotient at school age and increased risk of cerebral palsy [14]

In univariate analysis, odds of IVH significantly increased with decrease in GA, though this trend did not persist on multivariate analysis. A similar trend was reported in a retrospective study from Saudi Arabia [15]. Interestingly, preterm neonates with ELBW, weighing less than 1000g, exhibited slightly lower risk (aOR=6.3, 95%CI=1.7-23.1) of IVH in comparison to those with VLBW, weighing between 1000g to less than 1500g, (aOR=7.2, 95%CI=2.1-24.8). This observation may align with findings from another study that reported a negative association between IVH occurrence and small for gestational age (SGA), where presence of SGA was hypothesized to confer protection against IVH [16]. Collectively, these findings point towards the need to determine the potential neuroprotective mechanisms against IVH. Presence of RDS or PPROM had a two-fold significant increased risk of IVH occurrence. While RDS is a well-established risk factor, the role of PPROM remains inconclusive, with some studies reporting no significant association to IVH [17,18]. Surfactant administration which mitigates RDS, may contribute to elevated IVH incidence [19]. This association was counteracted on univariate analysis, which demonstrated a protective effect of surfactant therapy on IVH (cOR=0.6, 95%CI=0.3-0.9).

Paradoxically, preterm neonates with cyanosis (aOR=0.4, 95%CI=0.1-0.9) and hypothermia (aOR= 0.4, 95%CI=0.2-0.6) had a lower risk of IVH. Both cyanosis and hypothermia are known to be significantly associated with IVH. For instance, in a meta-analysis, hypothermic VLBW infants had an elevated risk of IVH (OR=1.9, 95%CI=1.0-3.1). It has been documented in an implementation research study that prevention of hypothermia may lower incidence of IVH [20,21]. Additionally, data from a national cohort study, premature infants with severe congenital heart disease (CHD) had a markedly increased likelihood of developing IVH (OR=6.2, 95%CI=5.6-6.9) [22]. Our inverse observations may be accounted for by the relatively lower proportions of enrolled preterm neonates presenting with cyanosis or hypothermia.

### Strengths and limitations of the study

This study highlights the IVH rate and factors associated with IVH in Tanzania. It also underscores the potential for targeted interventions to address modifiable risk factors for IVH. However, due to resource constraints, our study did not follow up on the neurodevelopmental outcomes of the preterm neonates with IVH. Furthermore, multi-center studies may be necessary to broaden the generalizability of our single center findings.

## CONCLUSION

There is still a significant IVH burden with low grade IVH predominating in Tanzania. Modifiable risk factors associated with IVH included RDS and PPROM which can be potentially be targeted with interventions to reduce IVH.

## Data Availability

All data produced in the present study are available upon reasonable request to the authors

## Ethical consideration

Informed written consent for enrollment into the study and open access publication was obtained from the neonates’ parents or guardians. Approval to conduct this study was sought and granted from the joint CUHAS/BMC Research and Ethics Committee (certificate number CREC/757/2024). Refusal to enroll in the study did not alter or jeopardize management. Participants’ confidentiality was strictly maintained, and data anonymized during and after the study.

## Data Availability

The de-identified participant data, data sets generated and analyzed are available from the corresponding author upon a reasonable and ethical request.

## Acknowledgments

We thank the study participants for their participation and contribution into the study. We also appreciate members of staff from the Departments of Paediatrics and Child Health as well as Radiology for their technical support during this study.

